# Craniofacial features of 3q29 deletion syndrome: application of next generation phenotyping technology

**DOI:** 10.1101/2020.09.18.20197665

**Authors:** Bryan Mak, Rossana Sanchez Russo, Michael J. Gambello, Emily Black, Elizabeth Leslie, Melissa M. Murphy, The Emory 3q29 Project, Jennifer Mulle

## Abstract

**Introduction:** 3q29 deletion syndrome (3q29del) is a recurrent deletion syndrome associated with neuropsychiatric disorders and congenital anomalies. Dysmorphic facial features have been described but not systematically characterized. This study aims to detail the 3q29del craniofacial phenotype and use a machine learning approach to categorize individuals with 3q29del through analysis of 2D photos.

**Methods:** Detailed dysmorphology exam and 2D facial photos were ascertained from 31 individuals with 3q29del. Photos were used to train the next generation phenotyping platform Face2Gene (FDNA, Inc, Boston, MA) to distinguish 3q29del cases from controls, using a proprietary algorithm. Area under the curve of receiver operating characteristic curves (AUC-ROC) were used to determine the capacity of Face2Gene to identify 3q29del cases against controls.

**Results:** In this cohort, the most common observed craniofacial features were prominent forehead (48.4%), prominent nose tip (35.5%), and thin upper lip vermillion (25.8%). The FDNA technology showed an ability to distinguish cases from controls with an AUC-ROC value of 0.873 (p = 0.006).

**Conclusion:** This study found a recognizable facial pattern in 3q29del, as observed by trained clinical geneticists and next generation phenotyping technology. These results expand the potential application of automated technology such as FDNA in identifying rare genetic syndromes, even when facial dysmorphology is subtle.

## Introduction

3q29 deletion syndrome is caused by a recurrent 1.6 megabase (Mb) deletion with an estimated prevalence of 1/30,000 births (Stefansson et al, 2014). The deletion is often de novo though up to 30% of cases may be inherited (Cox & Butler, 2015). Details about the phenotypic spectrum of the syndrome are emerging from both case reports and series (Ballif, 2008; Cox & Butler, 2015) and systematic studies (Glassford et al, 2016; Sanchez Russo et al, 2020).

The developmental and neuropsychiatric phenotypes associated with 3q29 deletion syndrome are well recognized. A previous Emory 3q29 Project study by Glassford et al (2015) using self-reported data from 44 study participants observed that 98% of 3q29 deletion carriers had developmental delay, and 89% of deletion carriers were symptomatic within the first year of life with feeding problems and failure to gain weight. A high incidence of dental problems, recurrent ear infections, and gastrointestinal disorders was also reported (Glassford et al, 2015). Individuals with the 3q29 deletion are at a 40-fold increased risk for developing schizophrenia (Mulle et al, 2015) and are at increased risk for autism spectrum disorder (ASD) and social disability in even the absence of autism (Sanders et al, 2015; Pollack, 2019). These reports have included limited data about physical and craniofacial characteristics, highlighting the need for further delineation of dysmorphic features in the syndrome.

Detailed data about facial dysmorphology in patients with 3q29 deletion syndrome is sparse. Most case reports and case series have documented some craniofacial abnormalities from retrospective reviews. A case series of 14 participants with 3q29 deletion syndrome described 4 participants with high nasal bridge, 4 with abnormal ear morphology, and 5 with microcephaly (Ballif et al, 2008). In a registry-based study of 44 study individuals, self-report data revealed a high proportion of participants with dental abnormalities including wide spaced teeth and dental crowding (Glassford et al, 2015). To date, a distinguishing facial phenotype associated with 3q29 deletion is not well-described in the medical literature.

The use of microarrays and next-generation phenotyping technology in the clinical setting by both genetics and non-genetics providers is increasing (Michelson & Clark, 2020; Zarate et al, 2019). As one example, the Face2Gene (FDNA Inc, MA, USA) platform uses deep learning technologies to identify facial phenotypes in rare disorders. In addition to clinical data, this platform provides a differential diagnosis that can aid in the diagnosis of various genetic disorders. In the context of 3q29 syndrome, if a subtle, but recognizable, facial phenotype exists and can be detected by next-generation phenotyping technology, it could be leveraged in the clinic to streamline genetic testing and identify individuals with this, shortening the time to diagnosis and allowing patients faster access to tailored interventions.

We therefore sought to delineate the craniofacial features of 3q29 deletion syndrome and ask whether next-generation phenotyping technology could be used to identify a characteristic facial dysmorphology associated with 3q29 deletion syndrome. At the present time, 3q29 deletion is uncharacterized by Face2Gene, thus the present research study is the first to train the system to recognize this copy number variant (CNV). This study will allow us to add to the growing body of knowledge about 3q29 deletion syndrome and to evaluate the clinical application of next-generation phenotyping technology.

## Methods

Study participant recruitment followed the criteria outlined in the study protocol for the Emory 3q29 Project (Murphy et al, 2018). Study participants were recruited from the 3q29 Deletion Registry (3q29deletion.org), a voluntary registry database housed at Emory University (Glassford et al, 2015). Eligibility criteria were as follows: age of at least six years, molecular diagnosis of 3q29 deletion syndrome, English fluency, and ability to travel to Emory University (Atlanta, GA, USA) for a full evaluation (Murphy et al, 2018). One exception to the age criterion was made; a 4.85 year old who was part of a previously-described multiplex family was included in the study (Murphy et al, 2020). At the evaluation, a trained clinical geneticist obtained a medical history and completed a detailed physical examination with attention to craniofacial features. 2D frontal and side photos were obtained during the physical examination.

After the visit, two clinical geneticists who conducted the medical history and physical exam discussed and evaluated all captured clinical data and 2D facial photographs for each participant. To standardize this process, each dysmorphic feature captured by the clinical geneticists was mapped to its associated Human Phenotype Ontology (HPO) term (Köhler et al, 2018), the standardized terms for phenotypic abnormalities. Descriptive statistics were used to describe the frequency of each phenotypic abnormality.

Face2Gene has two independent applications: the clinical application, which leverages the Face2Gene database to provide up to 30 diagnoses based on craniofacial features, and a research application, that can be used to identify the presence of distinct facial patterns in a syndromic cohort not yet characterized by Face2Gene. We uploaded a total of 31 2D frontal photos, one for each participant in our cohort, into the research application in a de-identified manner. Using the proprietary DeepGestalt algorithm described by Gurovich et al (2019), a mathematical representation, the descriptor, of the face was created for 3q29 syndrome. This facial descriptor can be shown as a two-dimensional mask or composite image. Moreover, this mathematical descriptor can be readily compared to other such descriptors for other syndromes or sets of participant photos. This comparison can be visualized as a graphical heatmap to show the degree of similarity between the two facial descriptors being compared.

The descriptor from the 2D facial photos of our subjects were then compared to an equally sized age-, ethnicity-, and gender-matched control set comprised of typically developing individuals with no known genetic syndromes provided by FDNA. To protect anonymity of the control cohort, only the composite photo can be viewed. Both the 3q29 deletion cohort and control cohorts are randomly divided into two sets, a training set, used to train the algorithm, and a test set, used to test the training. This random split is repeated ten times and the results are provided as mean and aggregate results in the form of the area under a receiver operating characteristic curve (AUC-ROC curve). The results of the binary comparisons are reported both numerically and graphically. A standard deviation is reported for the AUC and a p-value is calculated for the aggregate results from the ROC and score distribution. We performed four comparisons: all cases (n = 31) to all controls (n = 31), female cases (n = 11) to female controls (n=11), male cases (n = 20) to male controls (n = 20), and female cases (n = 11) to male cases (n = 20) as a negative control.

## Results

### Demographics

Demographic features for 31 study participants can be seen in **Table 1**. 20 (65%) were male; ages ranged from 4 - 39 years (median = 11 years). The cohort was stratified into four age brackets used by Face2Gene (stata in years: 4-6, 7-12, 13-18, 19-40). All four age brackets were represented in our cohort with most participants aged between 7 years and 12 years of age (11 participants, 35.5%). 28 (90.3%) of participants identified as Caucasian. The remaining 3 participants were denoted as mixed race, identifying as Caucasian and one other ethnicity.

**Table 1.** Demographics (N=31)

| **Table 1. Demographics (N=31)** |  |  |
| --- | --- | --- |
|  | **n** | **%** |
| **Sex** |  |  |
| Male | 20 | 64.5% |
| Female | 11 | 35.5% |
| **Age** |  |  |
| 4y-6y | 6 | 19.4% |
| 7y-12y | 11 | 35.5% |
| 13y-18y | 9 | 29.0% |
| 19y-40y | 5 | 16.1% |
| **Ethnicity** |  |  |
| Caucasian | 28 | 90.3% |
| Mixed Race* | 3 | 9.70% |

*Note.* \*Participants of mixed race identified as Caucasian and one other ethnicity.

### Dysmorphic Craniofacial Features

Results from our clinical analysis of observed craniofacial features and 2D is reported in Table 2. In each facial region, dysmorphic features were seen in the majority of study participants as follows: 70.9% of participants had at least one forehead or cranium abnormality, 56.8% had an eye or eyebrow abnormality, 74.2% had a nose or midface abnormality, 61.3% had a lip or oral abnormality, and 67.7% had a mandible or face shape abnormality. Within these regions, the most commonly seen dysmorphic features were prominent forehead (48.4%), wide nose (48.4%), and thin upper lip vermillion (25.8%). Low hanging columella (22.6%), prominent nasal bridge (22.6%), and incisor macrodontia (22.6%) were also common (Figure 1). Other observed features were rare and only observed once or twice, particularly other eye, mouth, and ear features.

**Table 2.** Most Commonly Described Craniofacial Features of 3q29 Deletion Cohort (N=31)

| HPO ID | HPO Term | Totals (n) | % (N=31) |
| --- | --- | --- | --- |
| HP:0000252 | Microcephaly | 5 | 16.1% |
| HP:0011220 | Prominent forehead | 15 | 48.4% |
| HP:0000582 | Upslanted palpebral fissure | 4 | 12.9% |
| HP:0000490 | Deeply set eyes | 7 | 22.6% |
| HP:0000368 | Low-set, posteriorly rotated ears | 4 | 12.9% |
| HP:0000272 | Malar flattening | 5 | 16.1% |
| HP:0000445 | Wide nose | 15 | 48.4% |
| HP:0000426 | Prominent nasal bridge | 7 | 22.6% |
| HP:0005274 | Prominent nasal tip | 11 | 35.5% |
| HP:0009765 | Low hanging columella | 7 | 22.6% |
| HP:0000322 | Short philtrum | 5 | 16.1% |
| HP:0000219 | Thin upper lip vermillion | 8 | 25.8% |
| HP:0011081 | Incisor macrodontia | 7 | 22.6% |
| HP:0000308 | Microretrognathia | 5 | 16.1% |
| HP:0000324 | Facial asymmetry | 5 | 16.1% |

### Face2Gene Analysis

Results from the Face2Gene analysis can be found in Figure 2 and Table 3. Group A consisted of all 2D frontal photos of the 31 participants in one cohort and the matched control cohort. The AUC-ROC curves (Figure 2) show that Face2Gene successfully differentiated between a 3q29 deletion participant and a control patient 87.3% of the time with a p-value of 0.006. Group B consisted of 20 male controls and 20 male cases. Similar to the full cohort comparison, Face2Gene was able to reach an AUC-ROC of 0.874 at a p-value of 0.016. Group C compared 11 female controls to 11 female 3q29 deletion participants. Group C did not meet significance (p=0.128) and could only differentiate between participants and controls 69.4% of the time. Lastly, group D compared the 3q29 deletion female cohort to the 3q29 deletion male cohort to see if there were discernable sex-attributed differences between the craniofacial phenotypes presented in females vs males. Group D did not meet significance (p=0.201) and could only differentiate between female and male cases 63.2% of the time.

**Table 3.** ROC Results for Whole Cohort and Gender Stratification

| **Table 3. ROC Results for Whole Cohort and Gender Stratification** |  |  |
| --- | --- | --- |
| Comparison Groups | AUC-ROC | p-value |
| All controls vs 3q29 Deletion Cases | 0.873 | 0.006 |
| Male controls vs. Male cases | 0.874 | 0.016 |
| Female controls vs. Female cases | 0.694 | 0.128 |
| Female cases vs Male cases | 0.632 | 0.201 |

**Figure 2.**
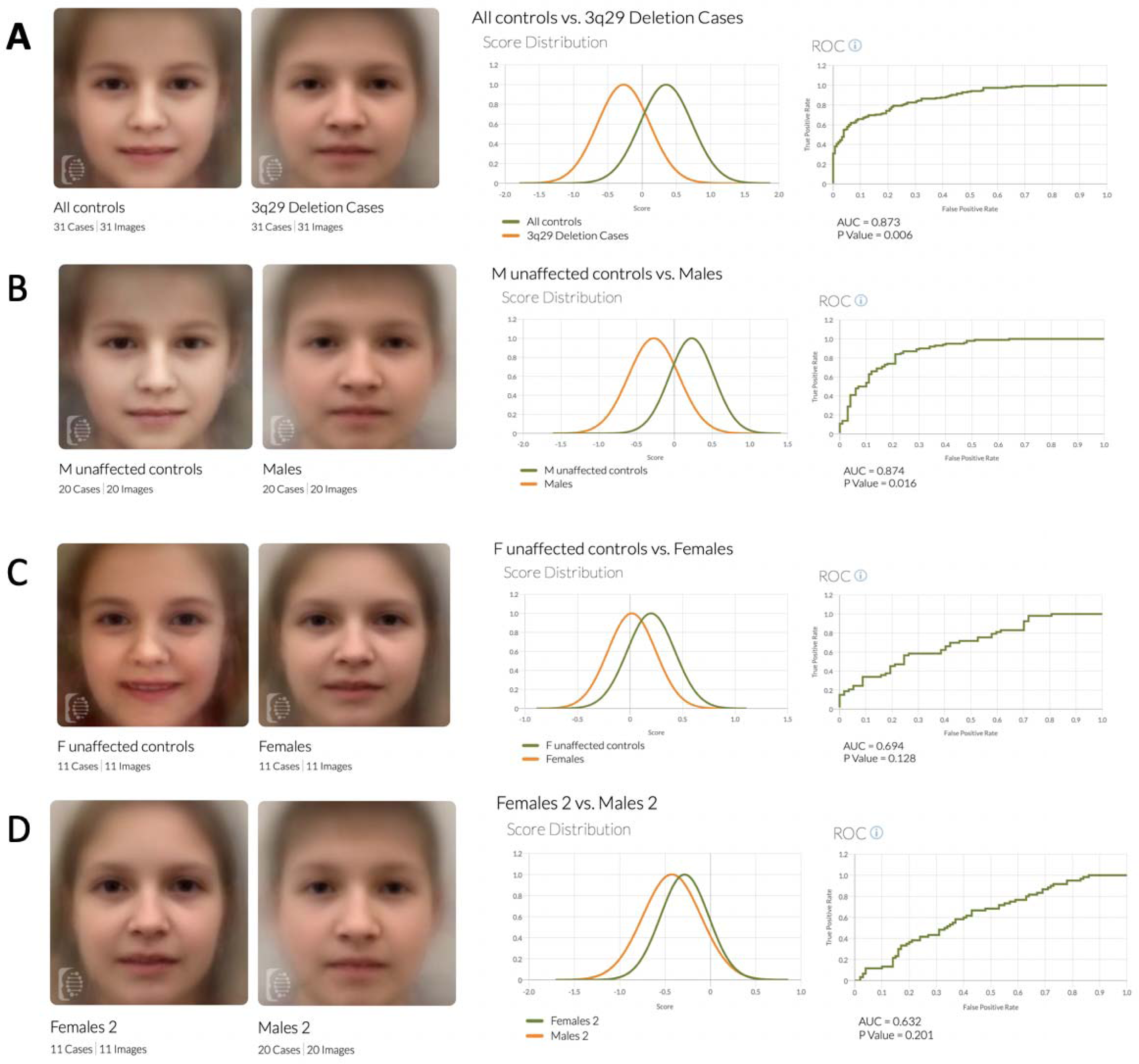
Face2Gene Composites and Binary Comparisons for Whole Cohort and by Gender *Note*. Comparison of control vs case composites and the binary comparison graphs. (A) Comparison of age-, gender-, and ethnicity-matched controls to the whole 3q29 deletion cohort. (B) Comparison of male controls to male cases. (C) Comparison of female controls to female cases. (D) Comparison of female cases to male cases.

### Common differentials

Face2Gene lists 30 differential diagnoses each participant independent of the research algorithm. Differentials are based off the binary descriptor and listed as high, medium, or low matches. How high a participant matches to a syndrome depending on how similar the participant’s face matches Face2Gene’s facial model for that given syndrome. For 13 out of 31 patients, Marfan syndrome was listed as one of the top 3 differentials, making it the most common differential offered for 3q29 deletion syndrome cohort by Face2Gene. One participant was listed as a high match for Marfan syndrome, 9 participants were listed as medium matches for Marfan, and 3 participants were listed as low matches for Marfan based on gestalt alone.

## Discussion

This is the first and largest study to systematically describe the craniofacial phenotype of 3q29 deletion syndrome. The focus of this study on the craniofacial phenotype builds upon and refines previously reported data. A literature summary of 40 previous case reports by Cox and Butler (2015), which includes a case series from Ballif et al (2008), and the previous 3q29 Project study by Glassford et al (2016) reported craniofacial findings that overlap with our study participants. Although our description of the 3q29 facial phenotypes is similar to previous studies, a notable difference are the frequencies at which some features were observed. For example, Ballif et al (2008) reported on 14 individuals with 3q29 deletion syndrome and found 72% with high nasal bridge 33% with posteriorly rotated ears, 40% with short philtrum, and 46.7% with microcephaly. In contrast, we found none of these features in greater than 25% of our cohort. Our participant sample was larger than previous studies, likely making the observed frequencies in this study more representative of the true frequencies. In addition, previous studies were culled reviews of the literature and may not have used the same terms to describe similar features. This study explicitly looked at craniofacial features and used a standardized nomenclature. The true frequencies of these features may be somewhere between the observed frequencies of these different studies.

By documenting common features seen in 3q29 deletion participants, we are able to aid clinicians in recognizing this rare copy number variant. In our experience with the 3q29 project (Glassford et al (2016)) and in the Ballif et al (2008) study, patients with 3q29 deletion syndrome are often referred to genetics clinics primarily for developmental delays and autism diagnoses. Chromosomal microarray (CMA) is still the first-tier clinical diagnostic test in individuals with developmental delays, autism, or congenital anomalies including dysmorphic features (Miller at al, 2010). By following this tiered testing schema and recognizing the dysmorphic features of 3q29 deletion carriers, it may be possible for patients to obtain a molecular diagnosis in a timely manner and prior to the onset of some of the more severe neuropsychiatric phenotypes (Sanchez Russo et al, 2020). Since the phenotype may be too subtle and too variable for clinicians to easily recognize 3q29 deletion syndrome as may be the case for other syndromes with characteristic facial phenotypes such as Cornelia-de-Lange or Down syndrome, using next-generation phenotyping in primary care clinics may aid in the recognition of this rare disorder (Latorre-Pellicer et al, 2020).

Research cohorts for facial models should reach an AUC-ROC of 0.80 – 0.85 or more, as this is the range where results are both sensitive and specific enough to have clinical utility. In the non-stratified 3q29 deletion cohort, Face2Gene was able to reach an area under the curve of 0.873 (p-value = 0.006). AUC-ROC curves of 0.8 to 0.85 or more have been described in the literature for conditions that presently have facial models within Face2Gene, like mucolipidosis type IV, where the AUC is 0.822 (p < 0.01) (Pode-Shakked et al, 2020). When comparing individuals with mucolipidosis type IV to a control group made up of 100 other genetic syndromes, Face2Gene reached an AUC of 0.885 (p < 0.001) (Pode-Shakked et al, 2020). Thus, a facial model for 3q29 deletion syndrome is feasible within Face2Gene.

When stratifying by gender, comparing males in the 3q29 deletion cohort to their matched controls reached significance and an acceptable AUC of 0.874, which is similar to that of the entire cohort. However, comparison to the female participants to matched controls did not reach significance. Likewise, comparing male to female participants was not significant, suggesting that the craniofacial phenotype of 3q29 deletion syndrome is not sex-specific or sex-influenced. This lack of statistical significance in the female control vs female participant group and the female participant vs male participant group may be due the smaller number of female participants. The cohort had only 11 females as compared to 20 males. Future studies may benefit from having a study population is more equal in gender representation. The aggregation of additional cases will also improve clinical detection rates for 3q29 deletion syndrome as our study is also limited by a lack of racial and ethnic diversity and a wide age range, both of which impact the ability of the algorithm to describe a facial gestalt and apply it clinically.

Marfan syndrome was the most commonly offered differential within the Face2Gene “Clinic” application. Marfan syndrome was a top 3 differential for 13 out of 31 participants ranging from low to high facial match. While Marfan syndrome and 3q29 deletion are clinically distinct, but they share some dysmorphic facial features including deep-set eyes and malar flattening. Interestingly, previously published data show other overlapping features between 3q29 deletion syndrome and connective tissue disorders including myopia, scoliosis, chest cavity deformity, long/slender fingers, foot deformity, and ligamentous laxity (Cox & Butler, 2015; Sanchez Russo et al, 2020). These data suggest the hypothesis that there may be an underlying mechanism within the 3q29 deletion syndrome leading to this connective tissue-like phenotype.

A limitation and consideration of our study is that our study is 90% Caucasian which has the ability to impact the Face2Gene facial recognition model as a whole. Males were disproportionately represented in our cohort which may have contributed to the stratified Face2Gene analysis not reaching significance. Lastly, our cohort had a very wide age gap with ages ranging between 4 – 40 years of age. This large age range may have an effect on clinical evaluation and the appearance of certain facial features. These limitations can all be ameliorated with a larger sample size of 3q29 deletion cases upon which to build the facial recognition model, which is a possible future direction. Regardless, this is the still the largest cohort of 3q29 deletion patients ascertained for systematic characterization of the 3q29 deletion facial phenotype.

## Conclusion

The medical community continues to move forward in better understanding 3q29 deletion syndrome. This study is the largest to systematically describe the craniofacial phenotype of 3q29 deletion syndrome. Our analyses suggest there may be a non-random pattern to the subtle facial dysmorphology associated with 3q29 deletion syndrome. This notion is supported by the application of machine-learning algorithms to 2D photos, where 3q29 deletion study subjects can be distinguished from control subjects with 87% accuracy from 2D photos alone. Future studies looking at the natural history of 3q29 deletion syndrome will provide invaluable information on how to better diagnose and treat these individuals.

## Data Availability

Due to the nature of this research, participants of this study did not agree for their data to be shared publicly, so supporting data is not available.

